# Cocooning is essential to relaxing social distancing

**DOI:** 10.1101/2020.05.03.20089920

**Authors:** Xutong Wang, Zhanwei Du, George Huang, Remy F Pasco, Spencer J Fox, Alison P Galvani, Michael Pignone, S. Claiborne Johnston, Lauren Ancel Meyers

## Abstract

As the first wave of COVID-19 recedes, policymakers are contemplating the relaxation of shelter-in-place orders. Using a model capturing high-risk populations and transmission rates estimated from hospitalization data, we find that postponing relaxation will only delay a second wave and cocooning vulnerable populations is needed to prevent overwhelming medical surges.

---

On December 31, 2019, the Wuhan municipal health commission first reported cases caused by the novel SARS-CoV-2 coronavirus (1). As of April 23, 2020, there have been over 2.5 million confirmed cases and more than 175,000 deaths across 210 countries (2). The US accounts for 31% of all confirmed COVID-19 cases and 23% of associated deaths worldwide (2). To mitigate the rapidly growing epidemics across the US,(3) the White House released guidelines on social distancing on March 16, 2020 (4), and over 42 states have since issued shelter-in-place orders, impacting more than 316 million people (5). While these measures have been effective in reducing disease spread, they also have had significant economic and social consequences.

On April 16, 2020, the White House issued the *Opening Up America Again* guidelines for a three-phased relaxation of social distancing measures. (6) However, city and state leaders remain divided regarding the speed and extent with which to proceed (7). Notably, the ‘gating criteria’ and ‘preparedness responsibilities’ of the White House plan do not include provisions for protecting vulnerable populations, frequently referred to as cocooning, such as increasing staff and resources at long-term care facilities, incentivizing individuals with high-risk comorbidities to remain at home, and enabling people experiencing homelessness to social distance (8). To quantify the life-saving importance of proactive cocooning of vulnerable populations, we have projected the impacts of relaxation with and without additional measures for vulnerable populations.

We built a granular mathematical model of COVID-19 spread in US cities that incorporates age-specific and risk-stratified heterogeneity in the transmission and severity of COVID-19 (Appendix Section 2) (9). We focus on the Austin-Round Rock Metropolitan Area (MSA) in the state of Texas (henceforth *Austin*), which is the fastest-growing large city in the US. We are providing daily decision support for city leaders and have access to individual-level data of COVID-19 hospitalizations, ventilator use, and deaths. Social distancing began in Austin when the public schools closed on March 14, 2020 and ramped up when the city enacted the *Stay Home-Work Safe* order on March 24, 2020. Local hospitalization data through April 23, 2020 indicate that this order has reduced COVID-19 transmission by 95% [95% CI: 70%-100%].

If social distancing measures are completely relaxed on May 1, 2020, we estimate that COVID-19 hospitalizations would surpass Austin’s surge capacity of 3,440 beds in 29 days (95% CI: 19 - 44), on May 30th (Figure). Assuming instead that individual behavior and public health efforts continue to reduce transmission by 75% (relative to early March), hospital surge capacity would be reached after 109 days (95% CI: 72-184), on August 18th. Superimposing cocooning by maintaining a 95% reduction in transmission risk for Austin’s high risk population— 547,474 of 2,168,316 total MSA residents (Appendix Section 3)—would avoid hospital surge. Furthermore, cocooning would reduce cumulative COVID-19 hospitalizations by 64.6% and cumulative deaths by 74.7% (Table A1). We expect that cocooning would affect a similarly dramatic reduction in mortality in communities worldwide. Postponing the relaxation of shelter-in-place measures would not be expected to prevent a second pandemic wave (Fig. A1), but may buy more time to protect vulnerable populations.

Cities are likely to experience second waves of COVID-19 when social distancing orders are relaxed. Our model indicates that Austin must continue to reduce COVID-19 transmission by at least 85% beyond May 1 to avert overwhelming hospital capacity by the end of 2020 (Fig. A1). Therefore, the cocooning of older adults and individuals with known high risk conditions (8) is paramount to protecting the thousands of lives in Austin and millions worldwide. Consequently, sheltering of vulnerable populations should be added to the national ‘gating criteria’ *prior* to the relaxation of social distance. Cocooning should be resourced proactively, including programs that enable high-risk members of the workforce to self-isolate and concerted efforts to shelter residents of long term care facilities and populations experiencing homelessness, in which risks associated with large proportions of high-risk individuals are compounded by group living conditions that amplify COVID-19 transmission (8).

## Data Availability

Not applicable

## Acknowledgements

We acknowledge the support of NIH grants R01AI151176 and U01GM087719 and CDC contract 75D-301-19-C-05930 and the critical discussions and parameter guidance from Matthew Biggerstaff, Michael Johannson, and the FluCode network at the US CDC, Mayor Steven Adler of Austin, Texas, and the White House Coronavirus Task Force.

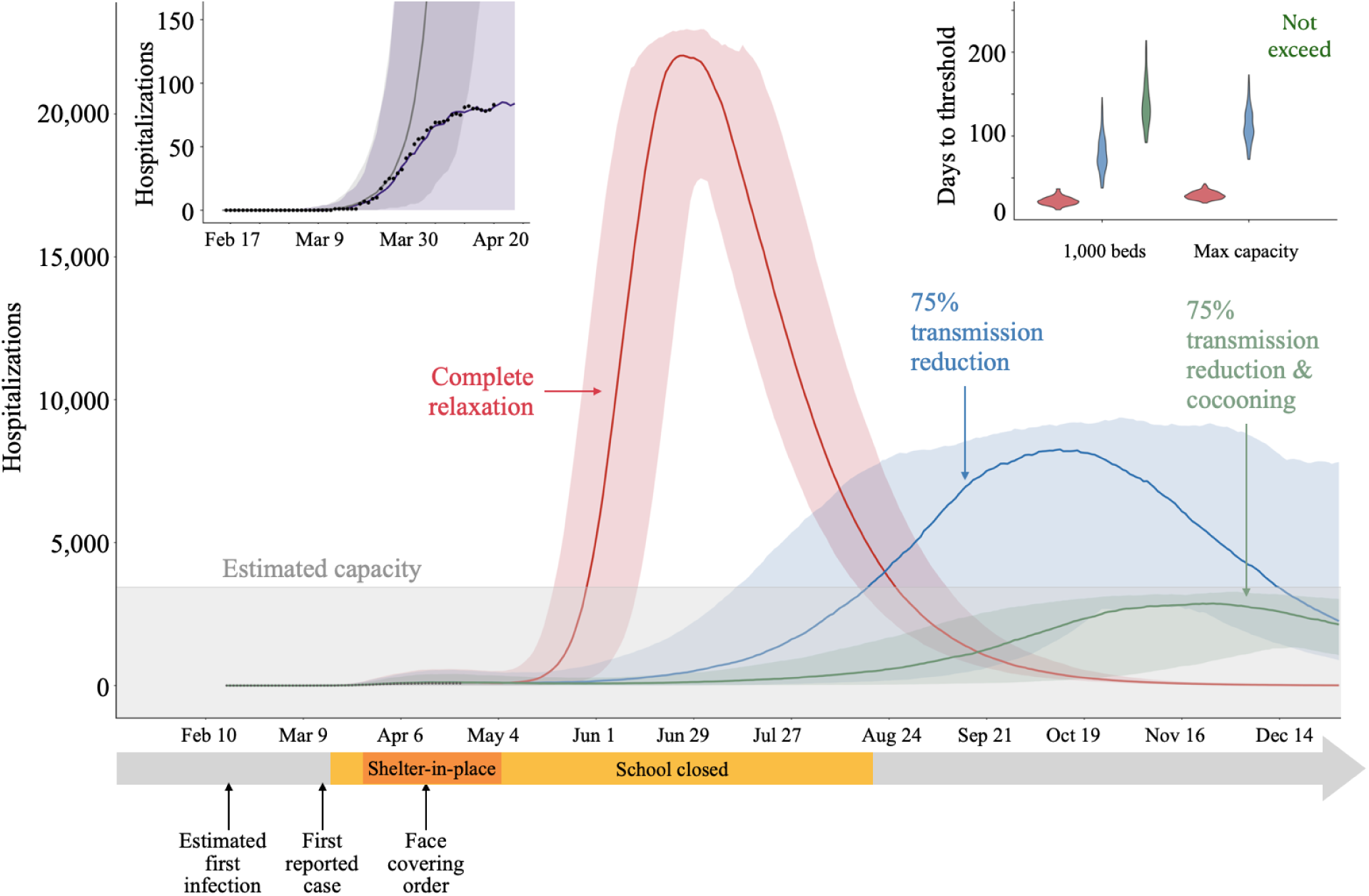
Figure. Projected daily COVID-19 hospitalizations in the Austin-Round Rock MSA from February 16 to December 31, 2020 assuming strict social distancing measures are relaxed on May 1, 2020. Upper left inset: These projections are based on a stochastic age- and risk-structured SEIR compartmental model of COVID-19 transmission fitted to comprehensive daily hospitalization data (black points) for the metropolitan area. The two curves are median projections based on the estimated transmission reduction of 95% [70%-100%] (purple) following the March 24, 2020 Stay Home-Work Safe order and a hypothetical scenario in which transmission was not reduced (gray). Main plot: Model fitting suggests that the ongoing COVID-19 epidemic in Austin was seeded by a local case on February 16, 2020; the first detected case was reported on March 13, 2020, schools were closed on March 15, and the shelter-in-place order was issued on March 24 and then amended to require cloth face coverings in public on April 13, 2020. The three projections assume that the order is relaxed on May 1st and schools open on August 18th. Following the May 1, we consider scenarios in which transmission rebounds to baseline (red), transmission is reduced by 75% through social distancing and aggressive testing, including in schools once the school year begins (blue), or transmission is reduced by 95% in high-risk populations through cocooning efforts and 75% in low-risk populations (green). Lines and shading indicate the median and 95% confidence interval across 200 stochastic simulations. The estimated total daily hospital capacity in the Austin-Round Rock MSA for COVID-19 patients is 80% of the 4,299 total beds (3,440), as indicated by gray shading along the bottom. Upper right inset: The time between the May 1st relaxation and COVID-19 hospital bed requirements exceeding either 1,000 or local capacity for each scenario, as indicated by corresponding colors.

